# Schistosomiasis in Humans, 1990-2041: Findings from the Global Burden of Disease 2021 Study and Predictions by Bayesian Age-Period-Cohort Analysis

**DOI:** 10.1101/2024.06.03.24308353

**Authors:** Qin Li, Yin-Long Li, Su-Ying Guo, Shi-Zhen Li, Qiang Wang, Wei-Na Lin, Li-Juan Zhang, Shi-Zhu Li, Xiao-Nong Zhou, Jing Xu

**Affiliations:** National Key Laboratory of Intelligent Tracking and Forecasting for Infectious Diseases, National Institute of Parasitic Diseases at Chinese Center for Disease Control and Prevention (Chinese Center for Tropical Diseases Research), NHC Key Laboratory of Parasite and Vector Biology, WHO Collaborating Centre for Tropical Diseases, National Center for International Research on Tropical Diseases, Shanghai 200025, China; School of Global Health, Chinese Center for Tropical Diseases Research, Shanghai Jiao Tong University School of Medicine, Shanghai 200025, China

**Author notes:** Corresponding author: Jing Xu. Email address: Qin Li;, Yin-Long Li;, Su-Ying Guo;, Shi-Zhen Li;, Qiang Wang;, Wei-Na Lin;, Li-Juan Zhang;, Shi-Zhu Li;, Xiao-Nong Zhou.

**Keywords:** Schistosomiasis, Disease burden, Trends, Bayesian Age-Period-Cohort model

## Abstract

**Background:** As the deadline for eliminating schistosomiasis approaches, more targeted and effective interventions should be proposed. We aimed to understand the burden among various gender, ages, countries, and continents and to analyze the trends in the burden of schistosomiasis from 1990 to 2041.

**Methods:** This study utilizes data from the Global Burden of Disease (GBD) 2021 to analyze the schistosomiasis burden trends from 1990 to 2021, including age-standardized rates of prevalence, Disability-Adjusted Life Years (DALYs), and death of different genders, ages, and regions. Data of schistosomiasis related anemia was also extracted and analyzed. Bayesian age-period-cohort (BAPC) models were used to assess and project the age standardized rates of prevalence, DALYs and death till 2041.

**Results:** Globally, the age-standardized rates of prevalence, DALYs, and death of schistosomiasis all present a declining trend. Individuals aged 15 to 29 years old present the highest age-standardized rates of prevalence and DALYs. The burden of schistosomiasis varies inversely with socio-economic development. The Years Lived with Disability (YLDs) rate for schistosomiasis-related anemia increases with the burden of schistosomiasis. The age-standardized rates of prevalence, DALYs, and death might tend to decline until 2041 in the world, Africa, Asia, and the Americas.

**Conclusions:** The burden of schistosomiasis is clustered in the 15 to 29 age group, which represents the strongest labor force. Additionally, reproductive-aged women also experience a significant disease burden. Targeted interventions including preventive chemotherapy, health behavior change, and communications should be proposed and covered this risk population.

**Author summary:** Schistosomiasis primarily affects more than 200 million people in Africa, Asia, and the Americas. To achieve the World Health Organization’s (WHO) 2030 target of eliminating schistosomiasis as a public health problem globally, understanding the distribution of schistosomiasis burden is crucial. In this study, we employed data from the Global Burden of Disease Study 2021 to analyze the burden of schistosomiasis across different regions, countries, genders, and age groups. Our analysis reveals that both the age standardized rates of prevalence and Disability-Adjusted Life Years (DALYs) of schistosomiasis reach the peak among individuals aged 15-29 years, who represent the highest potential for labor force participation and reproduction. The disease burden increases with the decline of Socio-demographic Index (SDI). However, countries with higher levels of healthcare level exhibit lower schistosomiasis-related anemia Years Lived with Disability (YLDs) rates. Most endemic areas in Asia and the Americas are supposed to achieve the target of schistosomiasis elimination before 2030, but Africa faces challenges in meeting it. Therefore, we advised endemic countries with lower SDIs to implement targeted interventions for the 15 to 29 age group. Meanwhile, improving healthcare level also be important to decrease the impact of schistosomiasis.

## Background

Schistosomiasis is a water-related infectious disease caused by a trematode of schistosomes, spreading through humans contacting with water contaminated by cercariae released by infected snails. In endemic areas, individuals may be repeatedly exposed to schistosomes from childhood or even fetal stages [1, 2], leading to chronic infections [3]. Such prolonged chronic infections can result in a range of complications, leading to irreversible damage to the gastrointestinal and urinary systems, as well as an increased risk of developing malignant tumors [4-6]. Additionally, schistosomiasis related anemia could lead to pregnant women getting negative reproduce outcome, such as low birth weight and premature birth in infants[7]. Infected infants and babies could experience higher rates of neonatal and perinatal mortality, as well as delayed child development, impacting their health throughout their lives[8].

Currently, schistosomiasis has spread to over 78 countries with nearly 800 million people at risk of infection [9]. It causes around 70 million disability-adjusted life years (DALYs) around world and impacts approximately 252 million people in tropical and subtropical regions [10-12], particularly in Africa, Asia, and the Americas [13, 14]. In 2020, WHO issued a roadmap for neglected tropical diseases and put forward the targets for schistosomiasis: (1) to eliminate schistosomiasis as a public health problem by 2030 in all endemic countries; (2) to interrupt the transmission of schistosomiasis in selected countries where transmission interruption is feasible [15]. To accelerate the process of schistosomiasis elimination, WHO launched a new guideline for control and elimination of human schistosomiasis in 2022 February based on evidence, recommending member states to extend preventive chemotherapy from school-aged children to all age groups at risk of schistosome infection [16]. However, as great heterogenicity existed in endemicity of schistosomiasis among population, communities and countries, and big gap existed between praziquantel donation and requirement, grasping the distribution of schistosomiasis burden across different regions, genders, and age groups, alongside analyzing long-term trend are crucial to support scientific policy-making, allocate resources appropriately, assess effectiveness of control programmes or guide intervention precisely.

The Global Burden of Disease (GBD) database, managed by the Institute of Health Measurement and Evaluation (IHME), encompasses a substantial panel of data spanning from 1990 to 2021, including the age-standardized rates of prevalence, Disability-Adjusted Life Years (DALYs), and death for schistosomiasis [17]. The objective of this study is to describe the distribution of schistosomiasis burden and schistosomiasis induced anemia, as well as to predict the epidemic trends of schistosomiasis for the next 20-years using the GBD 2021 data.

## Materials and Methods

### Data source

The data of age standardized rates of prevalence, DALYs, and death spanning from 1990 to 2021 were sourced from the official Global Burden of Disease (GBD) 2021 study website, which is openly available at http://ghdx.healthdata.org/gbd-results-tool. We selected all continents and countries around the world as geographic regions, specified ‘schistosomiasis’ as the cause, delineated ‘disability-adjusted life years (DALYs)’, ‘prevalence’, and ‘deaths’ as the chosen measurement indicators, and designed ‘Number’, ‘Rate’ as metric indicators within the database. In this research, we provided an analysis of schistosomiasis age standardized rates of prevalence, DALYs, and death across different regions. Furthermore, we also presented the changes in these indicators from 1990 to 2021, offering an overview of the evolving trends in schistosomiasis burden. Population estimates across different regions are sourced from the United Nations 2022 population projection data, which is disaggregated by year (up to 2100), age, and gender, and can be accessed at https://population.un.org/wpp/Download/Standard/Population/.

### Statistics

We employed numbers and rates along with 95% uncertainty intervals (UIs) to estimate the burden of schistosomiasis. The burden was assessed through prevalence, DALYs, and deaths, and was analyzed by genders, ages, locations, the age standardized rate of Years Lived with Disability (YLDs) of schistosomiasis induced anemia, and socio-demographic index (SDIs) by descriptive analysis. The prevalence number (or rate) denotes the count (or rate) of individuals with schistosomiasis within a population during a given year. DALYs are used to quantify the burden of disease, injuries, or health conditions caused by schistosomiasis on a population in this study. The number (or rate) of death refers to the count (or rate) of fatalities among individuals with schistosomiasis within a given year. SDI is employed to assess the socio-economic development level of countries or regions. YLDs measures the years of life lost due to premature death caused from schistosomiasis related anemia in this study.

For predicting the age standardized rates of prevalence, DALYs, and death from 2022 to 2041, we employed Bayesian age-period-cohort (BAPC) models. This model has demonstrated higher accuracy in predicting disease burden [18]. The predictions were conducted within an integrated nested Laplacian approximation framework using R packages BAPC and INLA. Throughout this study, we followed the Guidelines for Accurate and Transparent Health Estimates Reporting (GATHER) for cross-sectional studies [19]. All analyses were performed using R, version 4.3.1 (R Core Team, Auckland, New Zealand). Statistically significant results were defined as those with P-values *<* 0.05.

## Result

The changes of disease burden of schistosomiasis from 1990 to 2021. From 1990 to 2021, the trends of age-standardized rates of prevalence, DALYs, and deaths showed a decrease in all endemic countries. The global age standardized prevalence rate decreased by 0.23 per 100,000 (−0.33 to -0.13), the age standardized DALYs rate decreased by 0.41 per 100,000 (−0.52 to -0.32), and the age standardized death rate decreased by 0.67 per 100,000 (−0.70 to -0.63). (Fig1)

**Fig 1.**
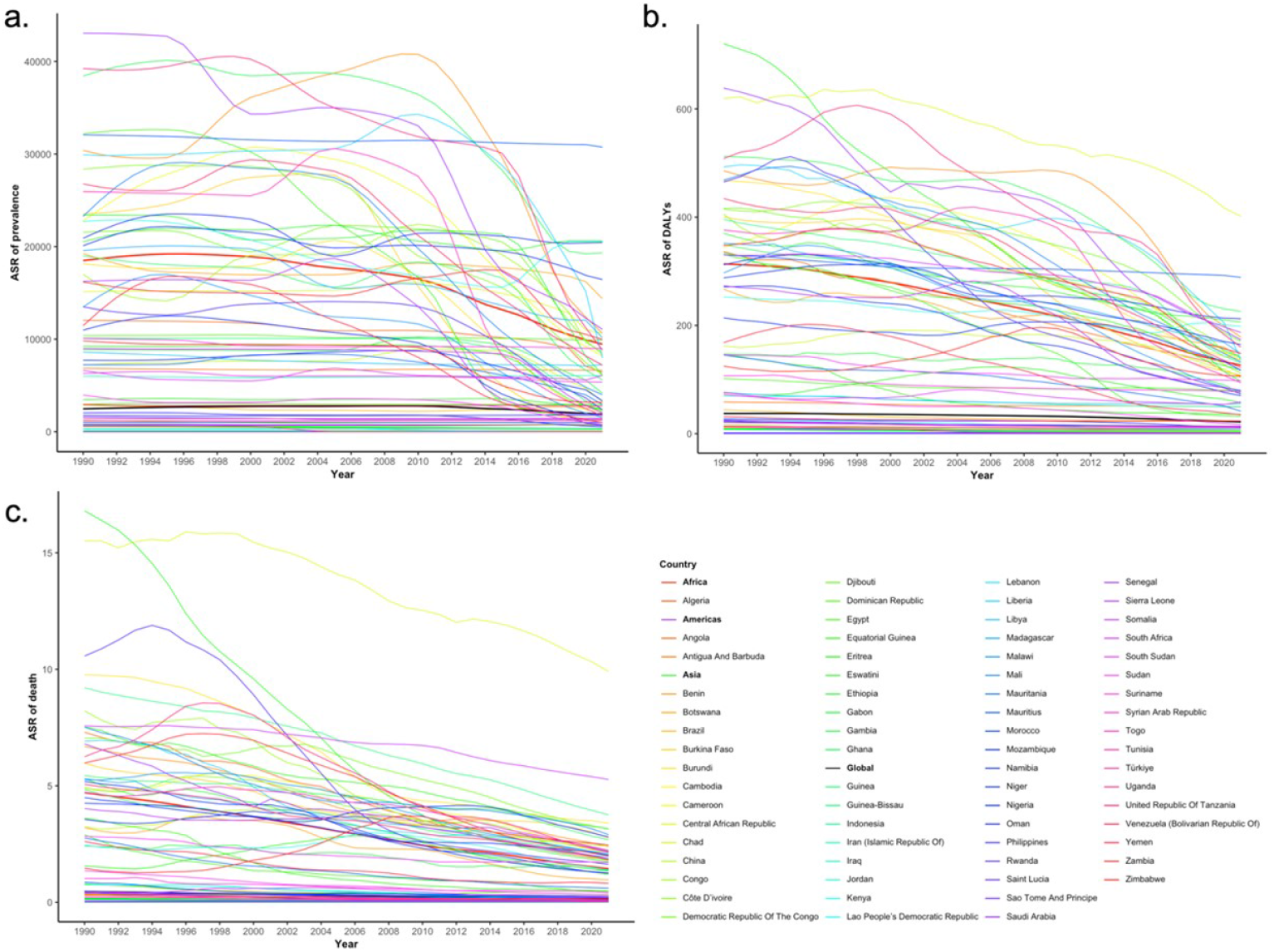
The changes of age standardized rates of prevalence, DALYs, and death across endemic countries from 1990 to 2021. a. Age standardized prevalence rate. b. Age standardized DALYs rate. c. Age standardized death rate.

In Africa, the age standardized prevalence rate decreased by 0.49 per 100,000 (−0.55 to -0.43), the age standardized DALYs rate decreased by 0.60 per 100,000 (−0.54 to -0.66), and the age standardized death rate decreased by 0.69 per 100,000 (−0.73 to -0.65). Among them, Egypt had the biggest decline in age standardized prevalence rate (−0.95 per 100,000, 95% UI: -0.96 to -0.92) and age standardized DALYs rate (−0.90 per 100,000, 95% UI: -0.92 to -0.88). Next were Burkina Faso (Age standardized prevalence rate: -0.94, 95% UI: -0.95 to -0.94; Age standardized DALYs rate: -0.73, 95% UI: -0.68 to -0.48; Age standardized death rate: -0.54 per 100,000, 95% UI: -0.67 to -0.37) and Rwanda (Age standardized prevalence rate: -0.82, 95% UI: -0.84 to -0.81; Age standardized DALYs rate: -0.84 per 100,000, 95% UI: -0.89 to -0.79; Age standardized death rate: -0.85 per 100,000, 95% UI: -0.90 to -0.76). (Fig1)

In Americas, the age standardized prevalence rate decreased by 0.05 per 100,000 (−0.12 to -0.03), the age standardized DALYs rate decreased by 0.35 per 100,000 (−0.48 to -0.24), and the age standardized death rate decreased by 0.64 per 100,000 (−0.67 to -0.61). The greatest declines were observed in Brazil (Age standardized prevalence rate: -0.13 per 100,000, 95% UI: -0.20 to -0.06; Age standardized DALYs rate: -0.48 per 100,000, 95% UI: -0.59 to -0.36; Age standardized death rate: -0.73 per 100,000, 95% UI: -0.76 to -0.71) and Suriname (Age standardized prevalence rate: -0.11 per 100,000, 95% UI: -0.29 to -0.08; Age standardized DALYs rate: -0.28 per 100,000, 95% UI: -0.49 to -0.07; Age standardized death rate: -0.59 per 100,000, 95% UI: -0.83 to 0.04). (Fig1) In Asia, the age standardized prevalence rate decreased by 0.48 per 100,000 (−0.53 to -0.43), the age standardized DALYs rate decreased by 0.67 per 100,000 (−0.75 to -0.59), and the age standardized death rate decreased by 0.85 per 100,000 (−0.88 to -0.83). The fastest decline in disease burden was observed in Turkey (Age standardized prevalence rate: -1.00 per 100,000, 95% UI: -1.00 to -1.00; Age standardized DALYs rate: -0.95 per 100,000, 95% UI: -0.97 to -0.93; Age standardized death rate: -0.92 per 100,000 95% UI: -0.95 to -0.88) and Iran (Age standardized prevalence rate: -0.98 per 100,000, 95% UI: -0.98 to -0.97; Age standardized DALYs rate: -0.84 per 100,000, 95% UI: -0.91 to -0.72; Age standardized death rate: -0.76 per 100,000, 95% UI: -0.89 to -0.44). (Fig1)

### The distribution of schistosomiasis burden in 2021

However, the global burden of schistosomiasis remained substantial now. Africa accounted for the majority of cases, comprising 84.25% (127,535,371.80/151,376,744.50) of the total population with schistosomiasis, followed by Asia (10.54%, 15,958,849.65/151,376,744.50) and the Americas (5.15%, 7,790,481.11/151,376,744.50). Schistosomiasis caused an estimated 1,746,333.31 DALYs globally (95% UI: 1,038,122.15-2,984,204.18), with Africa bearing the greatest burden at 87.92% (1,535,379.90/1,746,333.31), compared to Americas (4.12%, 71,901.42/1,746,333.31) and the Asia (7.88%, 137,607.03/1,746,333.31). The age standardized death rate was 0.15 per 100,000 (95% UI: 0.14-0.17) in 2021. Africa accounting for 87.28% (11,222.00/12,857.57) of schistosomiasis induced deaths, followed by Americas (4.69%, 602.15/12,857.57) and the Asia (7.83%, 1,006.30/12,857.57). (Fig 1)

In Africa, Ethiopia reported the highest age standardized prevalence rate at 20,629.50 per 100,000 (95% UI: 15,623.73-5,837.50), followed by Kenya (20,536.94 per 100,000, 95% UI: 15,062.14-26,748.39). Other significant countries included Nigeria (20,432.61 per 100,000, 95% UI: 14,906.45-26,836.63), and the Ghana (19,321.32 per 100,000, 95% UI: 13,477.54-26,646.31). The age standardized prevalence rate in the Americas was comparatively lower, with Dominican Republic (9,923.69 per 100,000, 95% UI: 6,620.53-14,219.80) and Suriname (9,002.65 per 100,000, 95% UI: 5,388.28-13,071.63).Asia had the lowest age standardized prevalence rate, with the Iraq and Oman at 6070.65 per 100,000 (95% UI: 4215.96-8609.98) and 1810.38 per 100,000 (95% UI: 1015.82-3070.71), respectively. (Fig 1)

Moving on to DALYs, in Africa, Central African Republic reported the highest age standardized DALYs rate at 282.88 per 100,000 (95% UI: 201.6-395.57), followed by Ghana (208.8 per 100,000, 95% UI: 110.78-377.10). Other significant countries included Nigeria (189.03 per 100,000, 95% UI: 99.99-344.13), and the Kenya (185.47 per 100,000, 95% UI: 93.53-355.39). The age standardized DALYs rate in the Americas was comparatively lower, with Suriname (80.19 per 100,000, 95% UI: 38.35-161.88) and Dominican Republic (82.38 per 100,000, 95% UI: 39.06-175.85). Asia had the lowest age standardized DALYs rate, with the Iraq and Yemen at 50.01 per 100,000 (95% UI: 23.25-104.51) and 35.16 per 100,000 (95% UI: 25.33-48.72), respectively. (Fig 1)

Finally, in terms of deaths, in Africa, Central African Republic reported the highest age standardized death rate at 9.91 per 100,000 (95% UI: 6.98-13.26), followed by Somalia (5.27 per 100,000, 95% UI: 3.74-7.05). other significant countries included Guinea-Bissau (3.75 per 100,000, 95% UI: 2.48-5.30) and the Burundi (3.39 per 100,000, 95% UI: 2.56-4.51). The age standardized death rate in the Americas was comparatively lower, with Suriname (0.42 per 100,000, 95% UI: 0.21-0.69) and Dominican Republic (0.32 per 100,000, 95% UI: 0.23-0.42). Asia had the lowest age standardized death rate, with the Yemen and Philippines at 0.81 per 100,000 (95% UI: 0.57-1.16) and 0.26 per 100,000 (95% UI: 0.22-0.30), respectively. (Fig 1)

### Burden by age and gender in 2021

Among different ages and genders, individuals aged 15 to 29 years showed the highest DALYs rate, with a prevalence rate of 11,059.93 per 100,000 (95% UI: 6137.15-16,707.66) and a DALYs rate of 104.79 per 100,000 (95% UI: 50.87-198.61). In contrast, the disease burden among children under 5 years old remained relatively low, with prevalence rate of 16.73 per 100,000 (95% UI: 3.73-49.96) and DALYs rate of 4.77 per 100,000 (95% UI: 3.05-6.51). The disease burden for males and females is comparable, both in prevalence and DALY rates.

Age standardized death rate increased with age, with the lowest rates observed among children aged 0 to 14 years old (0.04 per 100,000, 95% UI: 0.03-0.05). Notably, despite the higher age standardized prevalence rate and age standardized DALYs rate in the 15 to 29 age group, the age standardized death rate was relatively low (0.07 per 100,000, 95% UI: 0.055-0.085). Gender-specific age standardized death rate results indicated that both of women and men, aged 75 and older, have the highest age standardized death rate. (Fig 2)

**Fig 2.**
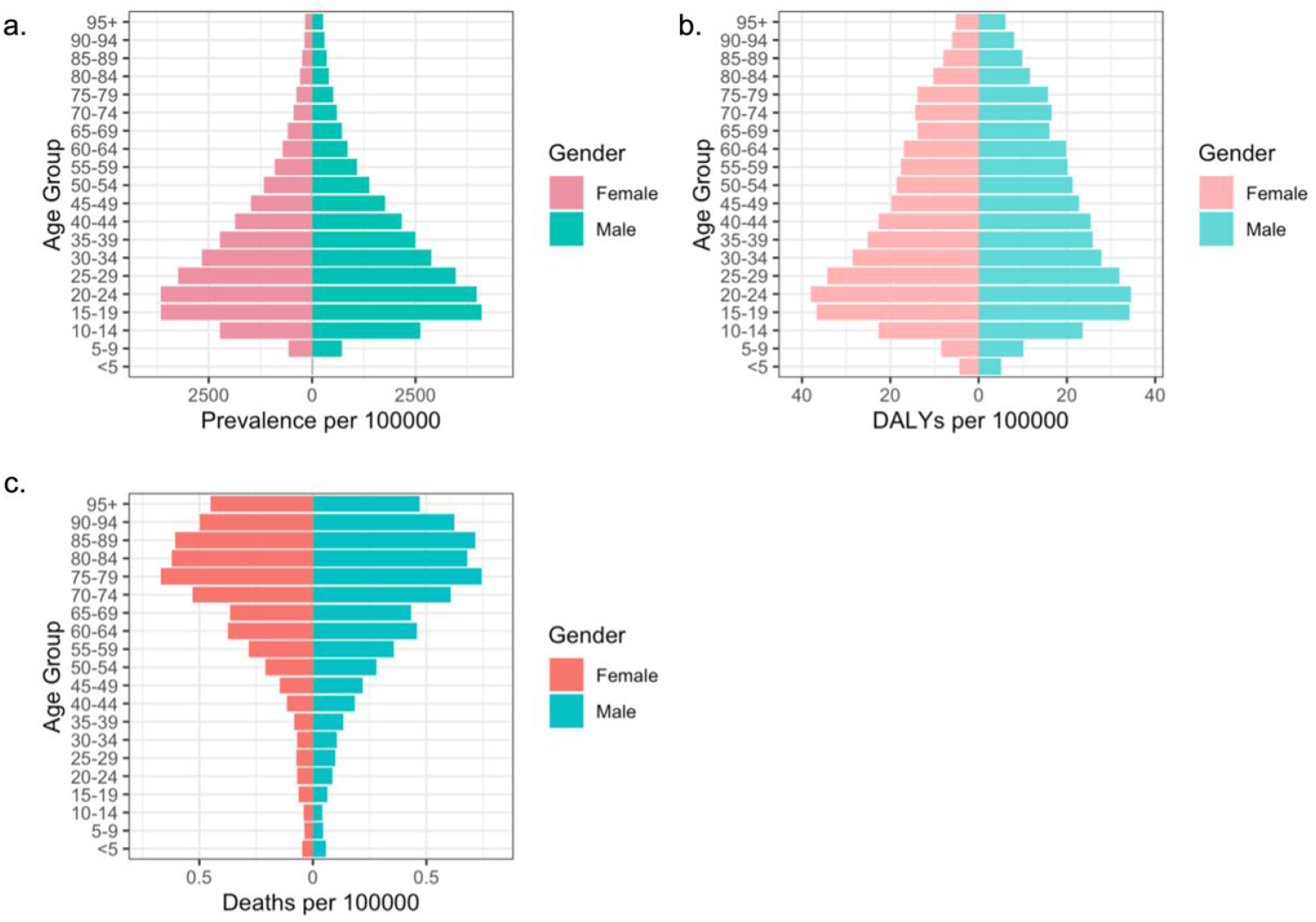
The prevalence, DALYs, and death rates across different genders and age groups in 2021 around the world. a. Prevalence rate. b. DALYs rate. c. Death rate.

### Burden variation by the socio-demographic index in 2021

The patterns of schistosomiasis burden vary significantly depending on the locations. Due to environmental limitations and specific habitat requirements of the intermediate host snails of schistosomes, some areas with lower SDI do not experience schistosomiasis endemicity. However, the age standardized rates of prevalence, DALYs, and death decrease as the SDI increases generally. Although the prevalence is somewhat lower, regions with the lowest SDI exhibit the highest age standardized DALYs rate and age standardized death rate. (Fig 3)

**Fig 3.**
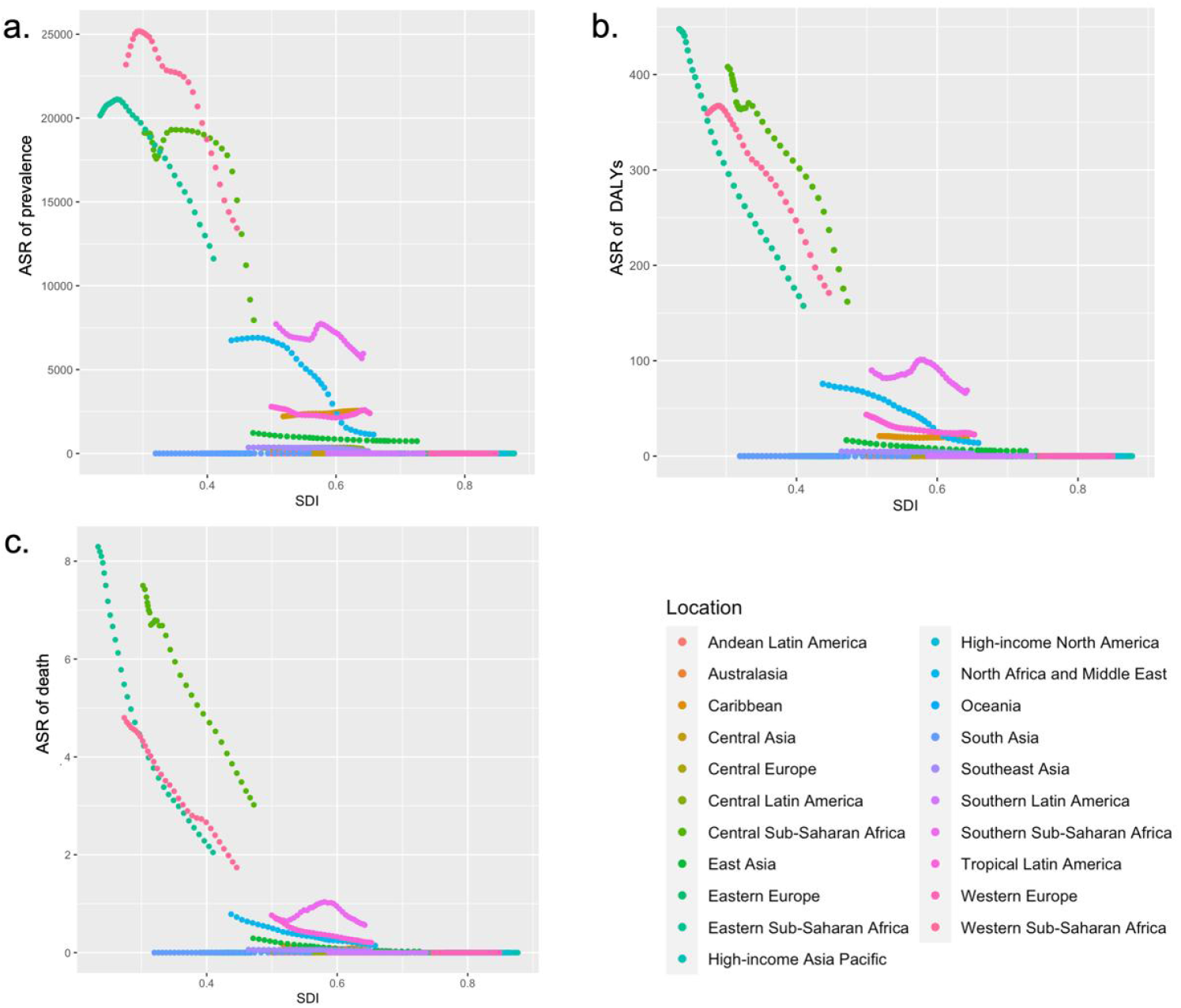
The age standardized rates of prevalence, DALYs, and death for GBD regions in 2021 by SDI. a. Age standardized prevalence rate. b. Age standardized DALYs rate. c. Age standardized death rate.

### Burden of schistosomiasis related anemia in 2021

Anemia is the significant symptom of schistosomiasis. In Africa, the age standardized Years Lived with Disability (YLDs) rate of schistosomiasis induced anemia in Nigeria leading with 46.34 per 100,000 (95% UI: 26.68-72.89), followed closely by Ghana (44.56 per 100,000, 95% UI: 24.00-72.58) and Senegal (33.80 per 100,000, 95% UI: 17.48-58.77); In the Americas, Suriname had the highest anemia YLDs rate at 12.82 per 100,000 (95% UI: 6.42-21.35) per 100,000, followed by the Dominican Republic (11.45 per 100,000, 95% UI: 6.05-19.89), then Antigua and Barbuda (7.51 per 100,000 95% UI: 1.64-16.33) and Brazil (3.16 per 100,000, 95% UI: 1.77-5.23); The age standardized YLDs rate of schistosomiasis-induced-anemia in Asia was notably lower, with the age standardized YLDs rate of Iraq at 6.35 per 100,000 (95% UI: 3.42-10.52), Syria at 2.09 per 100,000 (95% UI: 1.01-3.76), and Yemen at 2.00 per 100,000 (95% UI: 1.07-3.28). (Fig 4)

**Fig 4.**
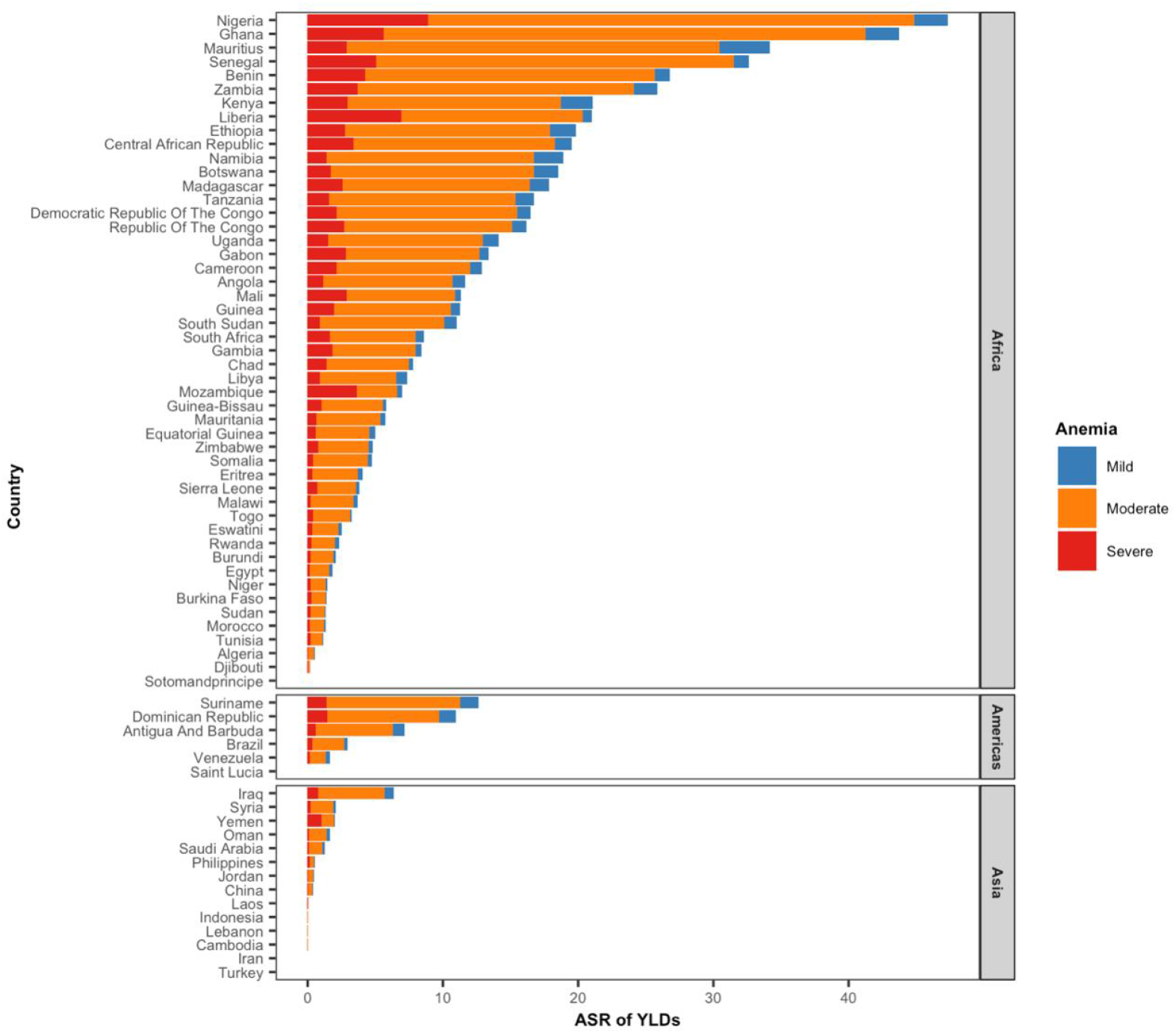
The composition of various levels anemia caused by schistosomiasis in endemic countries.

Globally, among different degrees of anemia, moderate anemia has the highest age standardized YLDs rate at 2.58 per 100,000 (95% UI: 1.48-4.07), followed by severe anemia with an age standardized YLDs rate of 0.53 per 100,000 (95% UI: 0.32-0.80) and mild anemia exhibits the lowest age standardized YLDs rate at only 0.22 per 100,000 (95%UI: 0.07-0.54). In Africa, anemia poses a particular severe impact, with the age standardized YLDs rate for moderate anemia reaching 17.55 per 100,000 (95% UI: 9.96-28.27), severe anemia at 2.82 per 100,000 (95% UI: 1.70-4.26), and mild anemia at 1.10 per 100,000 (95% UI: 0.36-2.65). The schistosomiasis-induced-anemia age standardized YLDs rate in the Americas is relatively milder, with a age standardized YLDs rate of 0.67 per 100,000 (95% UI: 0.37-1.11) for moderate anemia, 0.10 per 100,000 (95% UI: 0.06-0.17) for severe anemia, and 0.08 per 100,000 (95% UI: 0.03-0.19) for mild anemia, all lower than the global average. In Asia, the situation is mildest among them, with an age standardized YLDs rate of only 0.17 per 100,000 (95% UI: 0.10-0.28) for moderate anemia, while severe anemia and mild anemia have similar age standardized YLDs rates of 0.04 per 100,000 (95% UI: 0.02-0.05) and 0.03 per 100,000 (95% UI: 0.01-0.06) respectively. (Fig 4)

### Trends of schistosomiasis burden predicted by BAPC model from 2021 to 2041

Prediction results of BAPC model suggest that the global burden of schistosomiasis is expected to continue declining between 2021 and 2041 in Africa, Americas, and the Asia. By 2030, both Americas and the Asia are projected to meet the WHO’s schistosomiasis elimination standards (prevalence of schistosomiasis with heavy intensity in humans less than 1%). However, Africa is anticipated to struggle to meet these standards by 2030, with prevalence expected to remain above 1% after 2041. Despite this, by 2030, schistosomiasis-related disability and death in Africa are projected to decrease to extremely low levels, below 0.1%. (Fig 5)

**Fig 5.**
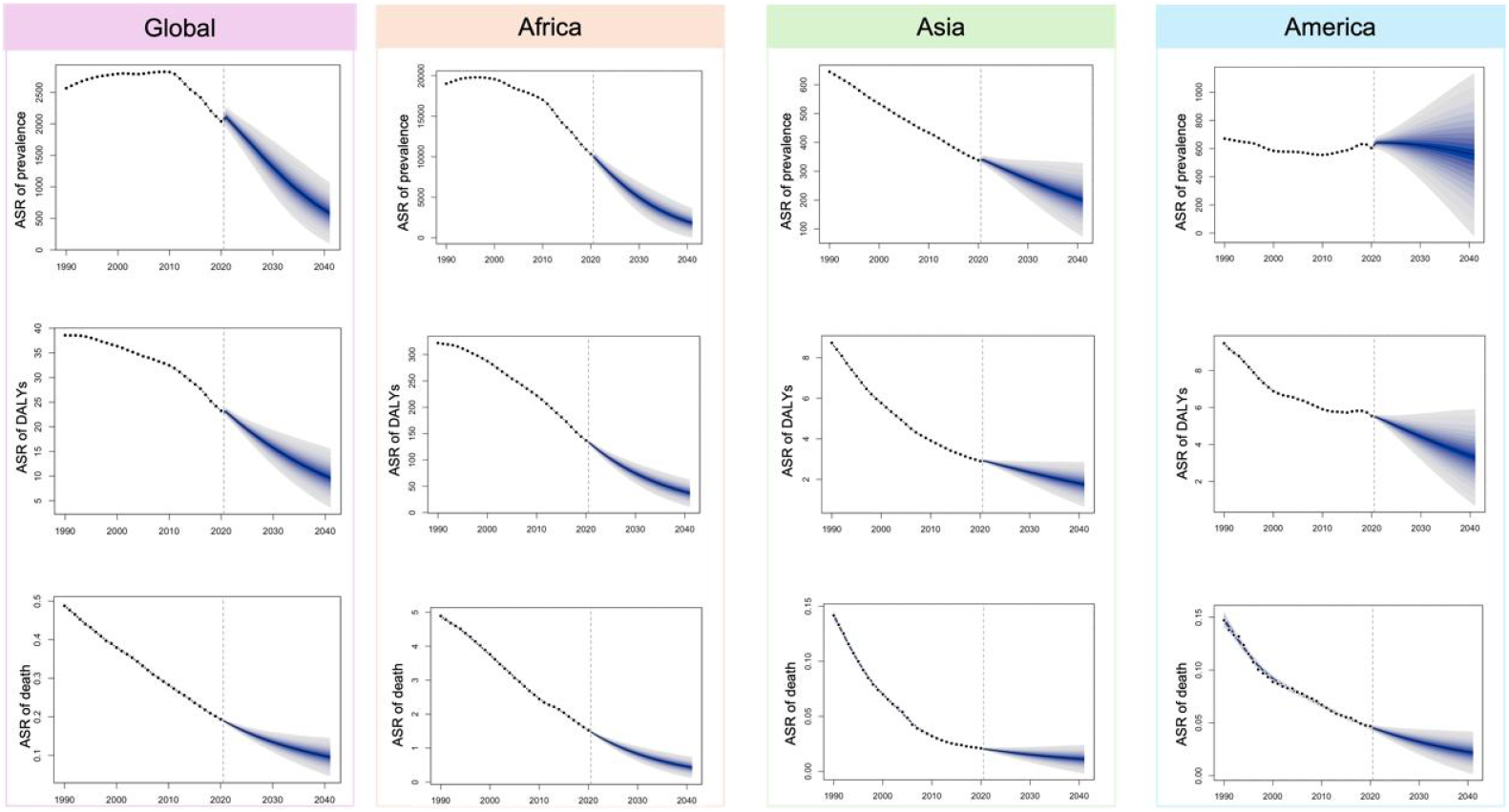
The change trends of the age standardized rates of prevalence, DALYs, and death from1990 to 2041 globally, as well as Africa, Americas, and the Asia.

## Discussion

This study offers a thorough analysis of the schistosomiasis burden by describing the age standardized rates of prevalence, DALYs, and death. Our findings indicate that from 1990 to 2021, there has been a declining trend in age standardized rates of prevalence, DALYs, and death globally, as well as in Africa, Americas, and the Asia. Meanwhile, the prediction result suggests this downward trend will continue from 2021 to 2041. It is anticipated that Americas and the Asia will reach the WHO’s schistosomiasis elimination targets by 2030, whereas Africa will need additional efforts to achieve similar goals. The broad geographic coverage and extensive temporal span of the study offer valuable insights into the global distribution and impact of schistosomiasis. Our results indicated that individuals aged 15 to 29, representing the strongest labor force, have the greatest disease burden. Combined with the results of correlation between schistosomiasis burden and SDI, it is evident that schistosomiasis-related labor loss constrains socioeconomic development in endemic areas, which, in turn, exacerbates the risk of infection. Meanwhile, among them, women, who are at childbearing age, have a notably high age standardized prevalence rate and age standardized DALYs rate. Previous research has revealed that pregnant women with schistosomiasis are three times more likely to suffer from anemia compared to non-pregnant individuals with the disease [7], severe schistosomiasis can cause negative pregnancy outcomes, including low birth weight, preterm birth, and infertility [20, 21]. Meanwhile, the infection during pregnancy not only heightens the susceptibility of pregnant women to schistosomiasis [22], but may also adversely affect the immune regulation of the fetus [23]. Given the economic disadvantaged regions always face a higher risk of schistosomiasis, interventions focused on the most productive labor force and children could offer meaningful contributions to addressing schistosomiasis.

As a tropical symptom of schistosomiasis, anemia contributes to a higher burden of various chronic diseases in endemic areas, particularly among the elderly [24]. Although anemia itself isn’t typically fatal, it leads to weakness and fatigue, thereby causing numerous often-overlooked losses. Nationally, countries with higher burden of schistosomiasis generally have higher age standardized YLDs rate for anemia, except for the Central African Republic, where the age standardized death rate is exceptionally high. Furthermore, in Yemen, Mozambique, Liberia, and three other three lower SDI countries with lower age standardized DALYs rates of schistosomiasis, over 50% of schistosomiasis-related anemia cases are severe, thereby different from the trend observed in other countries where moderate anemia predominates. This indicates that it is essential to propose healthcare measures tailored to low SDI countries [25, 26]. Therefore, efforts should be concentrated on enhancing healthcare capacity to reduce the schistosomiasis is urgent, particularly for low SDI countries [27].

The 2021 GBD data reveals that the burden of schistosomiasis in Americas and the Asia is lower than the global average, while Africa presents the highest burden. Combining with model results, there’s optimism that schistosomiasis elimination targets could be reached in endemic regions of Americas and the Asia before 2030. Therefore, it is advisable for the WHO to develop transmission assessment guidelines for countries with minimal schistosomiasis burden or nearing zero infection rates. Additionally, efforts for surveillance and consolidation are considered crucial for these countries[28]. Africa may require more time to reach the goal. The World Health Organization introduced new guidelines that extended the eligibility for preventive chemotherapy from primarily school-aged children to encompass all age groups [16]. With this new guideline, the actual situation may turn out to be even more favorable. Meanwhile, some researchers found that for people with multiple parasitic infections, more praziquantel needs to be given [29, 30]. Despite an increase in global donations of praziquantel over the past few years, resource inadequacy remains inevitable in comparison to the demand. Insufficient praziquantel availability may limit the expected outcomes. Given the factors such as poor sanitation, inadequate control of snails, limited health knowledge, and etc [31-35]. contribute to the heavier burden of schistosomiasis in these countries. Therefore, interventions related to water, sanitation, and hygiene (WASH), water engineering, focal snail control with molluscicides, and behavioral change should be emphasized, following WHO guidelines[16].

Our study also has other limitations. This study was based on the 2021 Global Burden of Disease (GBD) study, which includes data on schistosomiasis from around the world but does not specify the type of schistosomiasis. Therefore, it is impossible to analyze the global burden of different types of schistosomiases, which might be more helpful for policymakers. Additionally, while we used the SDI to describe socioeconomic differences among countries, further classification could provide further information for understanding burden of schistosomiasis.

## Acknowledgment

Thank you to the Global Burden of Disease 2021 team from the Institute for Health Metrics and Evaluation (IHME) for their invaluable contributions and dedication in compiling and managing the Global Burden of Disease database.

## Data availability statement

Burden of schistosomiasis from official Global Burden of Disease (GBD) 2021 study website http://ghdx.healthdata.org/gbd-results-tool. Population estimates across different regions are sourced from the United Nations 2022 population projection data https://population.un.org/wpp/Download/Standard/Population/.

## Authors’ contributions

Qin Li, Li-Juan Zhang, and Jing Xu developed the study protocol. Su-Ying Guo, Shi-Zhen Li, and Wei-Na Lin did data download. Qin Li, Yin-Long Li, and Qiang Wang did the data analysis. Qin Li drafted the report. Shi-Zhu Li, Xiao-Nong Zhou, and Jing Xu revised the report.

